# Prevalence and Prognostic Significance of Malnutrition Risk in Patients with Valvular Heart disease: Insight from the China-VHD Study

**DOI:** 10.1101/2023.05.15.23290021

**Authors:** Ziang Li, Bin Zhang, Zhe li, Yunqing Ye, Erli Zhang, Haitong Zhang, Qinghao Zhao, Zikai Yu, Weiwei Wang, Shuai Guo, Zhenya Duan, Junxing Lv, Bincheng Wang, Runlin Gao, Haiyan Xu, Yongjian Wu

**Author notes:** **Address for correspondence:** Haiyan Xu, Fuwai Hospital, National Center for Cardiovascular Disease, Chinese Academy of Medical Science and Peking Union Medical College, No.167 Beilishi Road, Beijing 100037, China., E-mail address., Yongjian Wu, Fuwai Hospital, National Center for Cardiovascular Disease, Chinese Academy of Medical Science and Peking Union Medical College, No.167 Beilishi Road, Beijing 100037, China. These authors contributed equally to the study and are joint first authors (Ziang Li, Bin Zhang). These authors contributed equally to the study and are joint corresponding authors.

## Abstract

**Background:** Previous studies on the prevalence and prognosis of nutritional status in valvular heart disease (VHD) were primarily limited to aortic valve stenosis. The nutritional status of other types of VHDs remained an underexplored area. This study aimed to evaluate the prevalence of malnutrition risk in different types of VHD and investigate the association between malnutrition risk and adverse clinical events.

**Methods:** A total of 8,908 patients with moderate or severe VHD in the China-VHD Study underwent echocardiography and malnutrition risk assessment using the geriatric nutritional risk index (GNRI). The primary outcome was 2-year all-cause mortality, while the secondary outcome was 2-year major adverse cardiovascular events (MACEs).

**Results:** Among 8,908 patients (mean age 61.8±13.4 years; 56% male), approximately half were at risk for malnutrition. Patients with moderate or severe malnutrition risk had significantly higher risks of all-cause death and MACEs in various VHDs compared to those without malnutrition risk (all p<0.02). The strongest association was detected in patients with tricuspid regurgitation (mortality: hazard ratio [HR], 3.18, 95% confidence interval (CI), 1.99-5.10, p<0.001; MACEs: HR, 2.33, 95% CI, 1.58-3.44, p<0.001). Adding GNRI score to the European System for Cardiac Operative Risk Evaluation improved risk stratification and outcome prediction (C-statistic p<0.001; likelihood ratio test p<0.001).

**Conclusion:** Malnutrition risk was prevalent in various VHDs and was strongly associated with increased mortality and MACEs. The GNRI score provided incremental prognostic information for clinical outcomes. Future research is needed to evaluate the impact of nutritional interventions on outcomes in these vulnerable patients.

**What is known:**

- Malnutrition is a prevalent condition among patients with aortic valve stenosis, and it represents a significant modifiable factor associated with adverse clinical outcomes.
- Nutritional screening tools is effective in identifying malnutrition in patients with aortic valve stenosis, but its usefulness for other types of valvular heart disease (VHD) is still unclear.

**What the study adds:**

-Malnutrition risk, as determined by geriatric nutritional risk index (GNRI), was prevalent in patients with all types of VHD.
- Compared with patients without malnutritional risk, those with moderate or severe malnutritional risk had a significantly increased risk of all-cause death and major adverse cardiovascular events (MACEs) in various types of VHDs, irrespective of body mass index and cardiac function.
- The addition of GNRI to the European System for Cardiac Operative Risk Evaluation (EuroSCORE II) amplified the stratification of patients at risk and improved outcome prediction.

## Introduction

The incidence of valvular heart disease (VHD) is on the rise in developing countries, primarily due to the high prevalence of rheumatic heart disease and the surge in degenerative valve disease.^1, 2^ Despite the medical advancements such as surgical or catheter interventions for valve replacement or repair, the death rate related to VHD has remained relatively stable since 2000.^1^ Therefore, it is crucial to identify high-risk patients based on modifiable clinical characteristics and intervene on these variables to reduce a patient’s risk.^2^

Recent evidence reported that malnutrition is a prevalent condition among patients with cardiovascular diseases, and it represents a significant modifiable factor associated with adverse clinical outcomes.^3–8^ Utilizing validated nutritional screening tools, such as the geriatric nutritional risk index (GNRI), prognostic nutritional index (PNI), or controlling nutritional status (CONUT), clinicians can easily identify malnutrition risk in patients with heart failure, stroke, or coronary artery disease,^5–8^ allowing for early intervention with personalized nutritional support and ultimately improving patient outcomes. However, there is a lack of comprehensive research on the prevalence and prognostic implications of malnutrition in VHD patients. Previous studies have shown that the GNRI is effective in identifying malnutrition in patients with severe aortic valve stenosis (AS)^4, 9^, but its usefulness for other types of VHD, such as aortic regurgitation (AR), mitral stenosis (MS), mitral regurgitation (MR), tricuspid regurgitation (TR), and multivalvular heart disease (MVHD), is still unclear. The latest guideline from the European Society of Cardiology recommends including nutritional assessment in the risk stratification of patients with VHDs.^10^ However, the current prognostic model for VHDs, such as the European System for Cardiac Operative Risk Evaluation II (EuroSCORE II) score, does not take nutritional status into account. ^11^ Therefore, further study is necessary to explore the prevalence, clinical associations, and prognostic consequences of malnutrition risk across various types of VHDs, as well as the potential benefits of integrating nutritional status into the EuroSCORE II model to improve risk stratification in VHD patients.

## Methods

The study adhered to the “Strengthening the Reporting of Observational Studies in Epidemiology” (STROBE) guideline and was approved by the Institutional Review Board at each center.^12^ All participants provided written informed consent prior to their inclusion in the study. The data are available from the corresponding author upon reasonable request.

### Study Population

The China Valvular Heart Disease (China-VHD, NCT03484806) is a nationwide, multicenter, observational, and prospective cohort study that focuses on patients with significant VHD. The enrollment was conducted between April and June 2018 at 46 medical centers across mainland China. A total of 13,917 consecutive patients with moderate or severe VHD, as defined by echocardiography using an integrative approach according to the 2014 American College of Cardiology/American Heart Association guidelines, were enrolled.^13^ Echocardiography images from the participating centers were evaluated for diagnostic accuracy and measurement consistency at the core laboratory in Fuwai Hospital before the recruitment, and the evaluators were blinded to the results obtained from individual centers. The study design and protocol have been previously published.^14^ For this analysis, we included 8,908 patients with native VHD of at least moderate severity who underwent echocardiography, physical measurement, and serum biochemical testing at baseline, after excluding those with previous valvular intervention, medical history of cancer, or infectious endocarditis. The study flowchart was presented in Figure S1.

### Malnutrition Evaluation

To assess the risk of malnutrition, the GNRI score was utilized, which was calculated using the following formula:1.489 × serum albumin (g/L) + (41.7 × weight [kg] / ideal body weight [kg]), where ideal body weight was determined using the Lorentz equations. ^4, 15^ For men, ideal body weight (kg) was calculated as body height (cm) – 100 – (body height - 150/4), while for women, it was calculated as body height (cm) – 100 – (body height - 150/2.5). A weight-to-ideal body weight ratio of 1 was assigned to patients whose body weight exceeded the ideal body weight.^6, 15^ Patients were further classified into four categories based on their level of malnourishment risk: no malnourishment risk (>98), low risk (92 to 98), moderate risk (82 to 91), or severe risk (<82). In subgroup analysis based on VHD type, we combined the moderate and severe risk groups due to the low number of subjects with severe risk. Body mass index (BMI) was calculated as weight (kg) divided by the square of the body height (in meters) and patients were classified as underweight (<18.5 kg/m^2^), normal weight (18.5 to 23.9 kg/m^2^), overweight (24.0 to 27.9 kg/m^2^), or obese (≥28 kg/m^2^) according to the recommendations of the Working Group on Obesity in China.^16^

### Clinical outcomes and Follow-up

The primary outcome was all-cause mortality, while the secondary outcome was major adverse cardiovascular events (MACEs), including cardiovascular death, hospital for heart failure, myocardial infarction, and stroke. Trained research fellows or coordinators conducted follow-up assessments at 0.5 year, 1 year, 1.5 years, and 2 years after VHD admission using face-to-face, telephone interviews, or medical records. Death and intervention reports were thoroughly scrutinized and validated by investigators at each participating center.

### Statistical Analyses

The mean ± standard deviation (SD) or median with interquartile range (IQR) were used to summarize continuous variables, depending on whether the distribution was normal or not. Categorical variables were expressed as counts and percentages. The differences between groups were evaluated using the one-way analysis of variance or Kruskal-Wallis test for continuous variables, and the chi-square test for categorical variables.

We initially investigated the relationship between GNRI and clinical outcomes by using restricted cubic splines. Kaplan-Meier curves were generated to visualize survival distributions, and log-rank tests were conducted to compare survival among various groups. Cox proportional hazards analyses were performed to compute hazard ratios (HRs) with 95% confidence intervals (CIs). To evaluate the prognostic impact of malnutrition, multivariate models were developed, which adjusted for variables associated with a poor prognosis based on clinical plausibility or p-value of <0.05 in univariate Cox analyses. The adjusted models included age, sex, BMI, cardiomyopathy, aortic disease, chronic obstructive pulmonary disease (COPD), pulmonary hypertension, New York Heart Association (NYHA) functional class >III, VHD severity, estimated glomerular filtration rate, left ventricular ejection fraction (LVEF), and valvular intervention.

Subgroup analyses were also performed, stratifying by age, gender, NYHA functional class, severity and etiology of VHD, and intervention strategy (intervention or conservative medical treatment). Participants were further categorized into four groups based on their BMI (high, ≥24 kg/m^2^; low, <24 kg/m^2^), LVEF (high, ≥50%; low, <50%), NT-proBNP (high, ≥1,000 pg/ml; low, <1,000 pg/ml), EuroSCORE II (high, ≥4%; low, <4%), and Geriatric Nutritional Risk Index (GNRI) (moderate or severe malnutrition risk <92; absent or mild malnutrition risk, ≥92), respectively. Kaplan-Meier curves were generated for each of the four combinations, and the log-rank test was used to compare the survival curves.

To assess the additional predictive value of GNRI, we included the dichotomized GNRI (moderate-severe malnutrition risk threshold) in a base model (EuroSCORE II) and compared their performance. Discrimination was evaluated by calculating the C-statistic, while calibration was assessed using the likelihood ratio test and Bayesian information criteria. All p-values were two-sided, and those below 0.05 were considered statistically significant. The analyses were performed using R version 3.5.2 (R Foundation for Statistical Computing, Vienna, Austria).

## Results

### Baseline Characteristics

Among the 8,908 patients, 4,971 (56%) were male and the mean age was 61.8±13.4 years. The median LVEF was 53.9%±13.1% and the median NT-proBNP was 1,361 pg/ml (IQR: 439.0, 3,851.3). Hypertension was the most common comorbid condition, presented in 44.6% of cases, and 40.0% of patients had dyspnea NYHA III or IV. The distribution of BMI categories was as follows: 581 patients (6.5%) were under weight, 4,433 (49.8%) were normal weight, 2,980 (33.5%) were overweight, and 914 (10.3%) were obese (Figure 1). Among the VHD subtypes, 465 (5.2%) had isolated AS, 1,241 (13.9%) had AR, 447 (5.0%) had MS, 2,182 (24.5%) had MR, 1,329 (14.9%) had TR, and 2,589 (29.1%) had MVHD. Table 1 summarizes the baseline characteristics of the overall cohort and the respective VHD subtypes.

**Figure 1.**
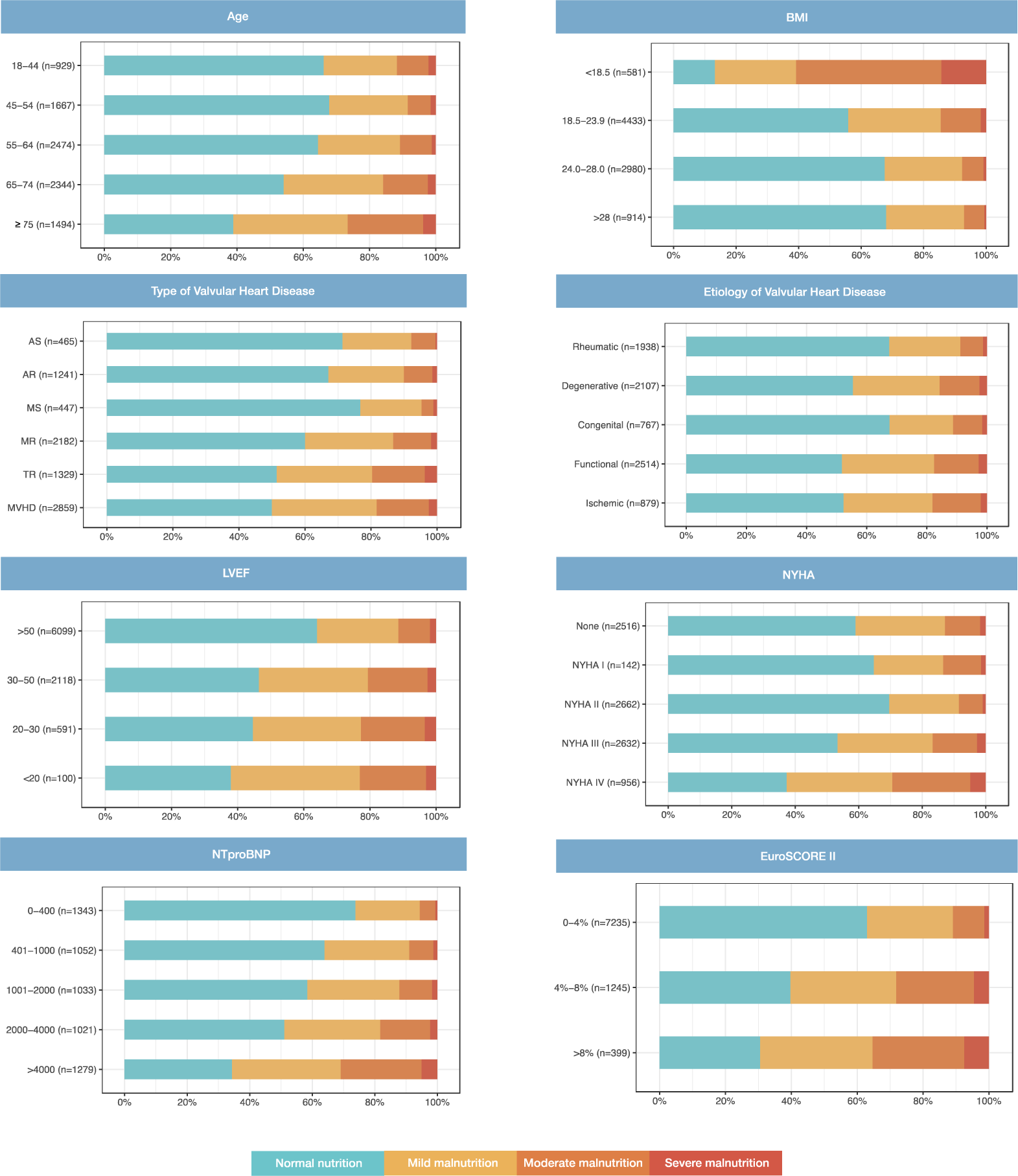
Prevalence of Malnutrition Risk in Different Subgroups. Prevalence of malnutrition risk in different subgroups of patients with valvular heart disease according the geriatric nutritional risk index. BMI, body mass index. LVEF, left ventricular ejection fraction. NYHA, New York Heart Association. EuroSCORE II, European System for Cardiac Operative Risk Evaluation II.

**Table 1.**
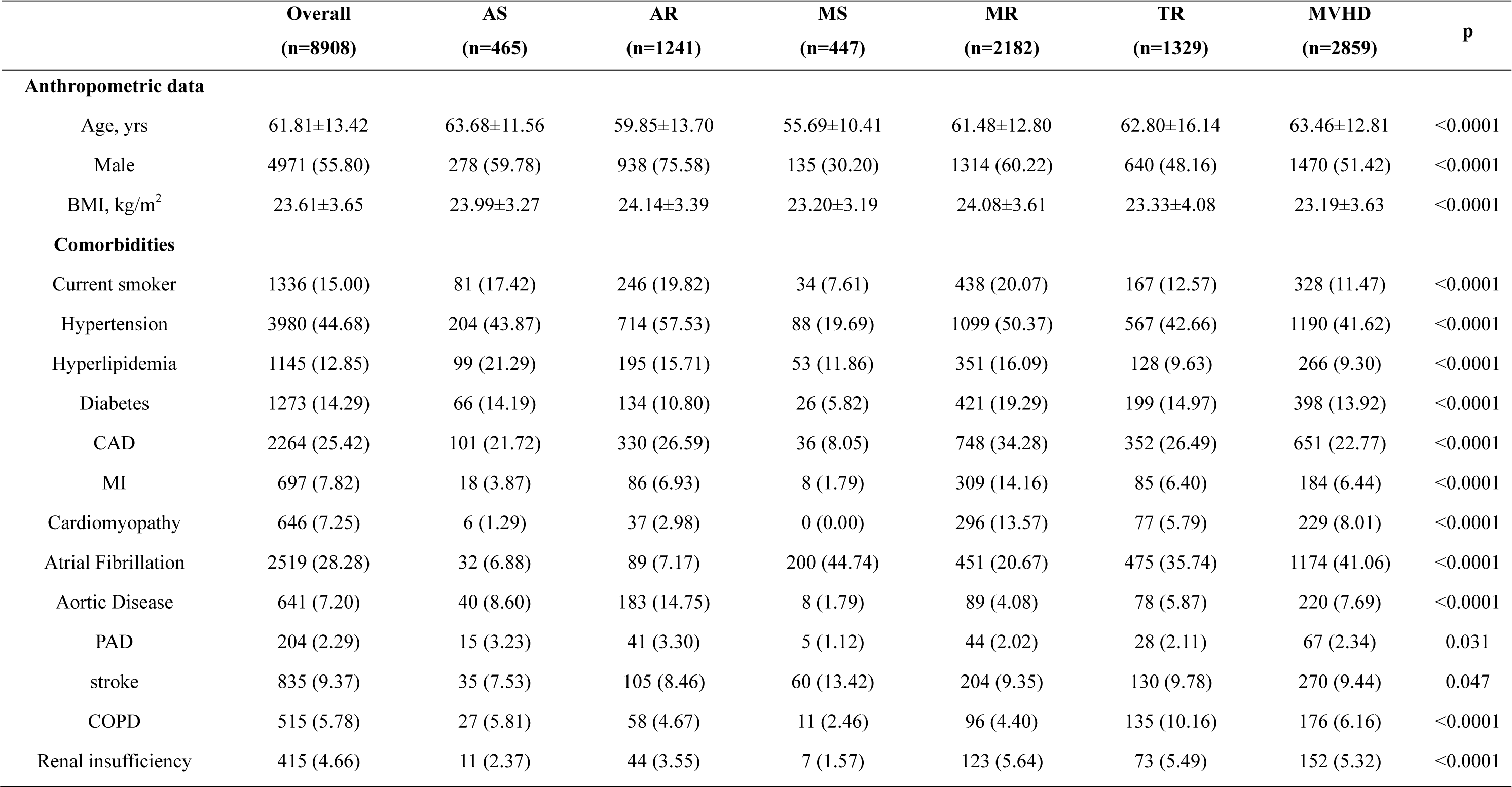
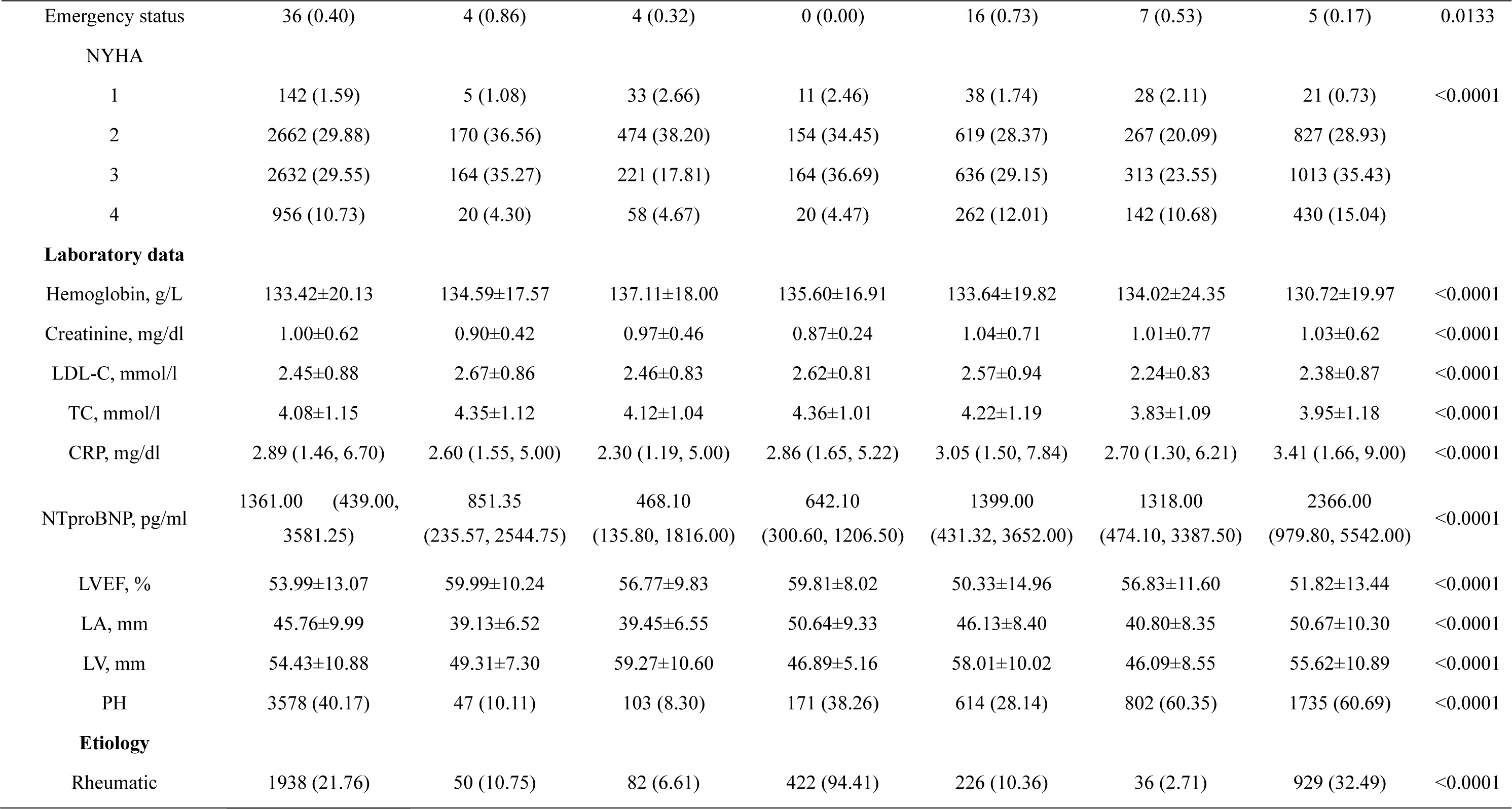
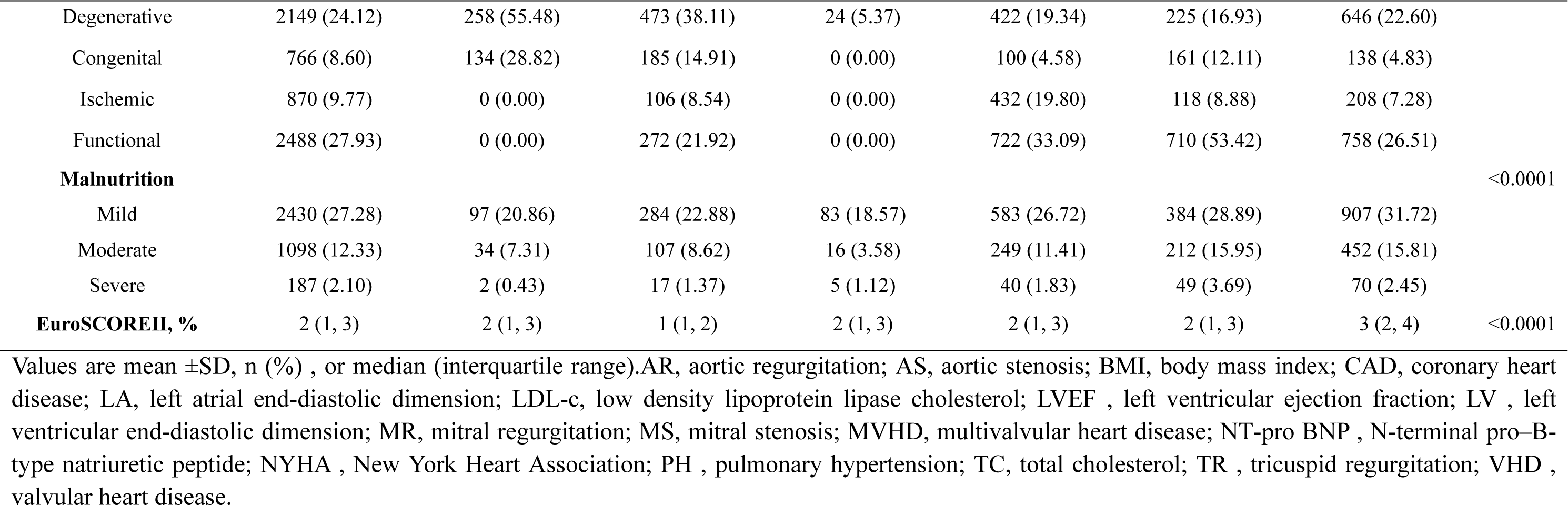
Baseline Characteristics of the study Population by Type of VHD

### Prevalence and Clinical Associations of Malnutrition

Based on GNRI calculation, 2,430 (27.2%), 1,098 (12.3%), and 187 (2.1%) patients were identified as having mild, moderate, and severe malnutrition risk, respectively. The proportion of individuals at risk of malnutrition did not differ significantly between young and middle-aged individuals, with rates of 33.8%, 32.2%, and 35.5% for ages 18-44, 45-54, and 55-64, respectively, while the proportion of individuals at risk of malnutrition was significantly higher in older age groups, with rates of 45.9% for ages 65-74 and 61.0% for ages 75 and above (Figure 1). In terms of types of VHD, patients with MVHD and TR had a higher prevalence of malnutrition risk (50.0% and 48.5%, respectively) compared to those with AS and MS (28.6% and 23.2%, respectively). With regards to the etiology of VHD, the proportion of malnutrition risk was relatively low in patients with rheumatic heart disease (32.5%) and congenital heart disease (32.3%) compared to patients with degenerative (44.5%), functional (48.2%), and ischemic heart disease (47.7%) (Figure 1). Besides, patients with malnutrition risk were more likely to have comorbidities such as diabetes, coronary artery disease, atrial fibrillation, stroke, COPD, worse NYHA class and NT-proBNP levels, compared to those with normal nutritional status (Table S1-7, Figure 1). Notably, while malnutrition was more common among patients who were older, had lower BMI, and poor cardiac function, the GNRI score identified a notable proportion of malnourished patients (30%) who were young and middle-aged (between 18-55 years), obese, or had low levels of NT-proBNP (<1000 pg/ml) or EuroSCORE II (<4%) (Figure 1).

### Follow-Up Data and Predictors of Clinical Events

At the 2-year follow-up, 702 patients (7.88%) died and 987 (11.08%) experienced MACEs. The overall survival rates at 1 and 2 years were 94.1%±0.5% and 91.7%±0.6%, respectively. The incidence rates for each cardiovascular event were as follows: 2.61% (n=465) for cardiovascular death, 2.76% (n=491) for heart failure, 0.24% (n=42) for myocardial infarction, and 0.57% (n=101) for stroke (Figure S1). Restricted cubic splines revealed curvilinear patterns linking GNRI and primary and secondary outcomes in various types of VHD. In the whole cohort, a curvilinear relationship between continuous GNRI scores and HRs of clinical outcomes was identified (Figure 2), with the curve increasing faster as GNRI scores fell below 98. All types of VHD showed a monotonic decrease in HRs with greater GNRI scores, and a remarkably sharper increase in HRs was observed for AR, MR, TR, and MVHD compared with AS and MS. In all types of VHD, patients with moderate to severe malnutrition risk had higher incidences of all-cause mortality and MACEs than those with mild or without malnutrition risk, regardless of whether the scores were used as a continuous or categorical variable (Figure 2-4).

**Figure 2.**
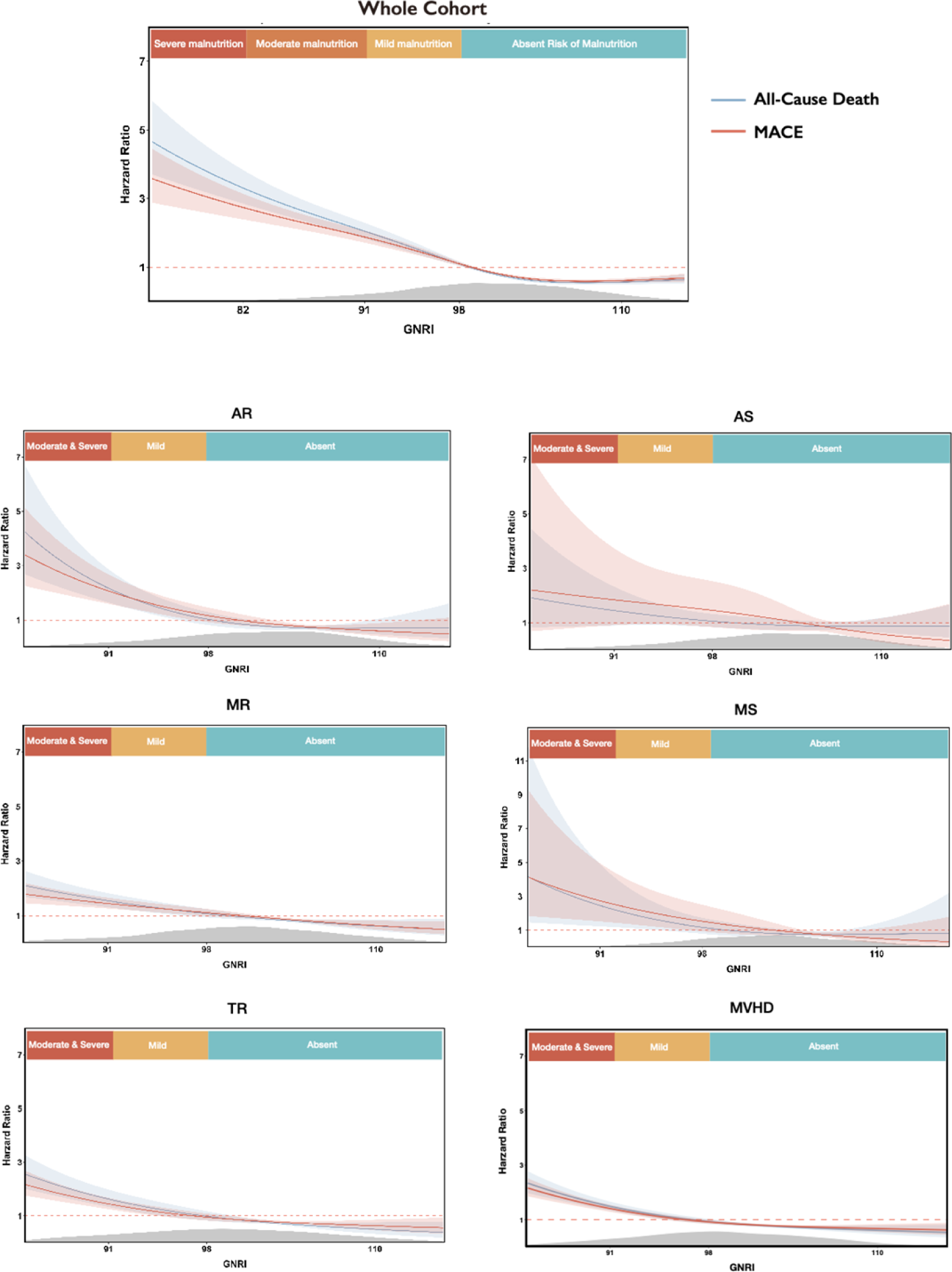
Impact of GNRI score on clinical outcomes in patients with Various VHDs. Restricted cubic splines demonstrated the shape and strength of association between GNRI score and clinical outcomes. The curves were presented with 95% confidence intervals. The gray area indicated the density of the populations. AR, aortic regurgitation. MS, mitral stenosis. MR, mitral regurgitation. TR, tricuspid regurgitation. MVHD, multivalvular heart disease.

**Figure 3.**
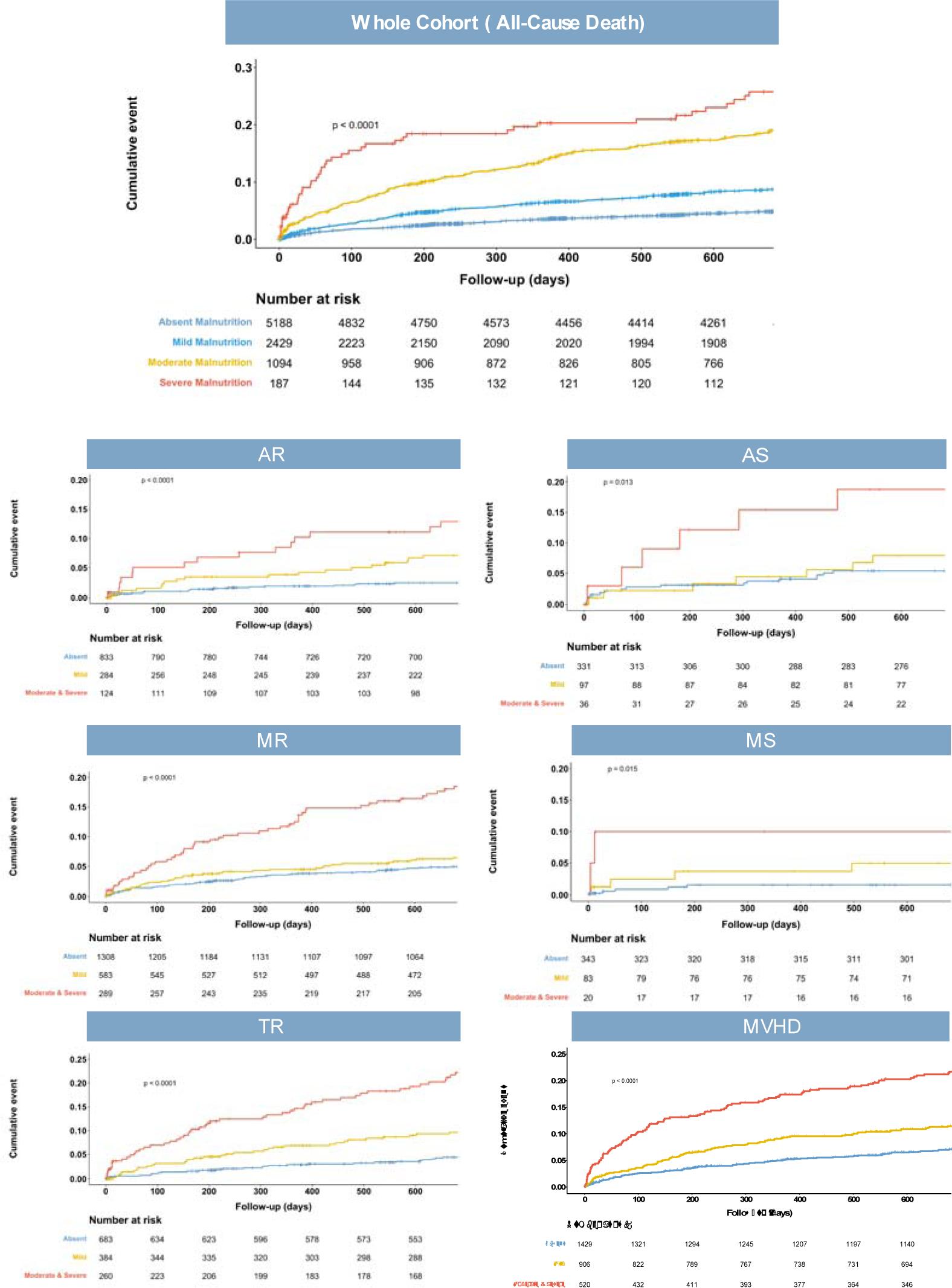
Malnutrition Degrees and Risk of Mortality. Abbreviation: AR, aortic regurgitation. MS, mitral stenosis. MR, mitral regurgitation. TR, tricuspid regurgitation. MVHD, multivalvular heart disease.

**Figure 4.**
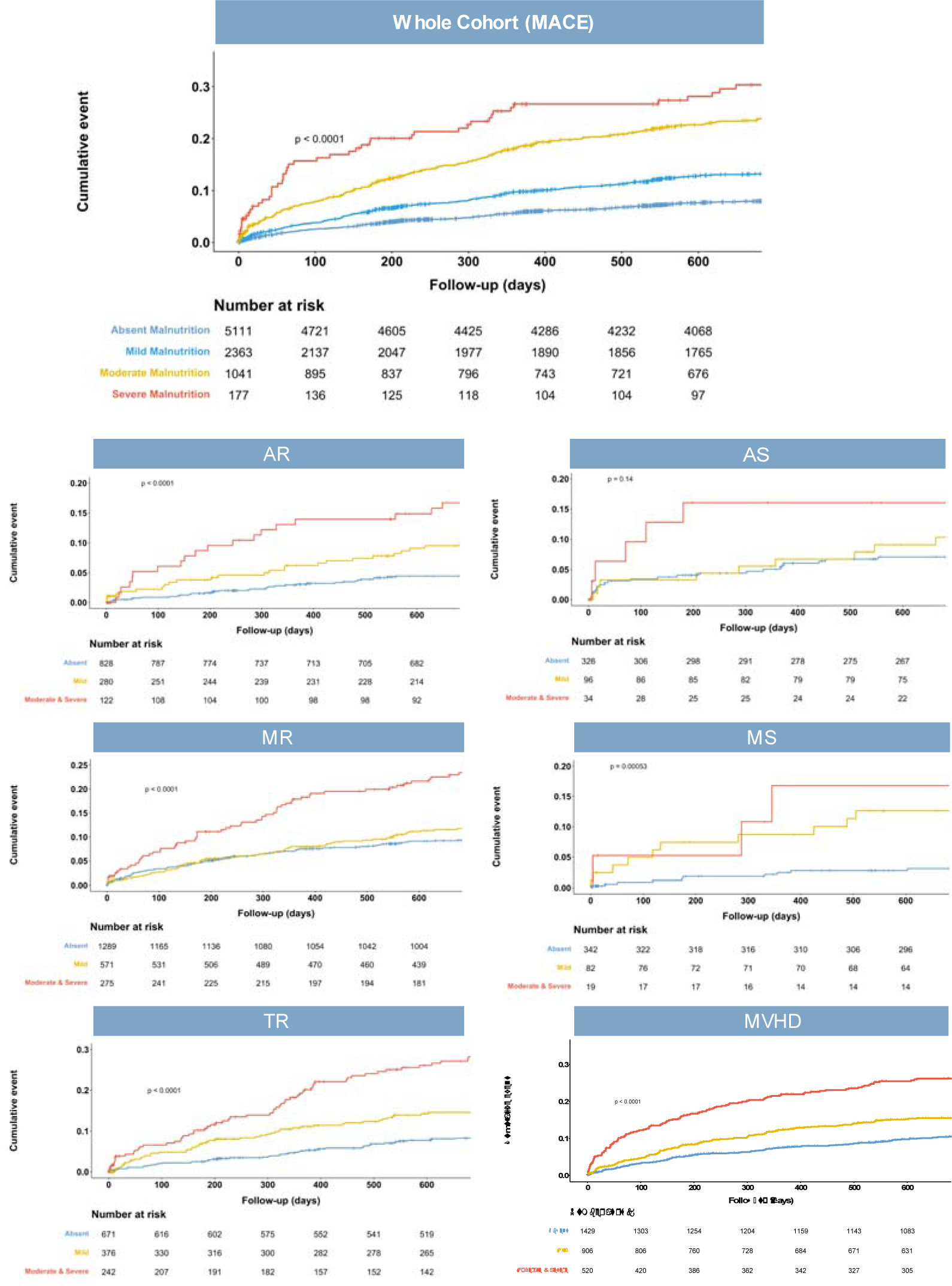
Risk of Mortality for Prespecified Groups Stratified by Nutritional status. Abbreviation: AR, aortic regurgitation. MS, mitral stenosis. MR, mitral regurgitation. TR, tricuspid regurgitation. MVHD, multivalvular heart disease.

Univariable analyses revealed that moderate to severe malnutrition risk was significantly associated with poor outcomes in all types of VHD, except for AS (Table 2, Table S8). After adjusting for age, sex, hypertension, coronary artery disease, atrial fibrillation, cardiomyopathy, COPD, pulmonary hypertension, creatinine, the severity of NYHA, valve intervention, and LVEF, the association between moderate to severe malnutrition risk and all-cause mortality and MACEs remained significant (Table 2). The association was found to be stronger in patients with TR (mortality: HR, 3.18,95% CI, 1.99-5.10, p<0.001; MACEs: HR, 2.33, 95% CI, 1.58-3.44, p<0.001) and MVHD (mortality: HR, 2.93,95% CI, 1.80-4.79, p<0.001; MACEs: HR, 2.37, 95 % CI, 1.48-3.80, p<0.001).

**Table 2.**
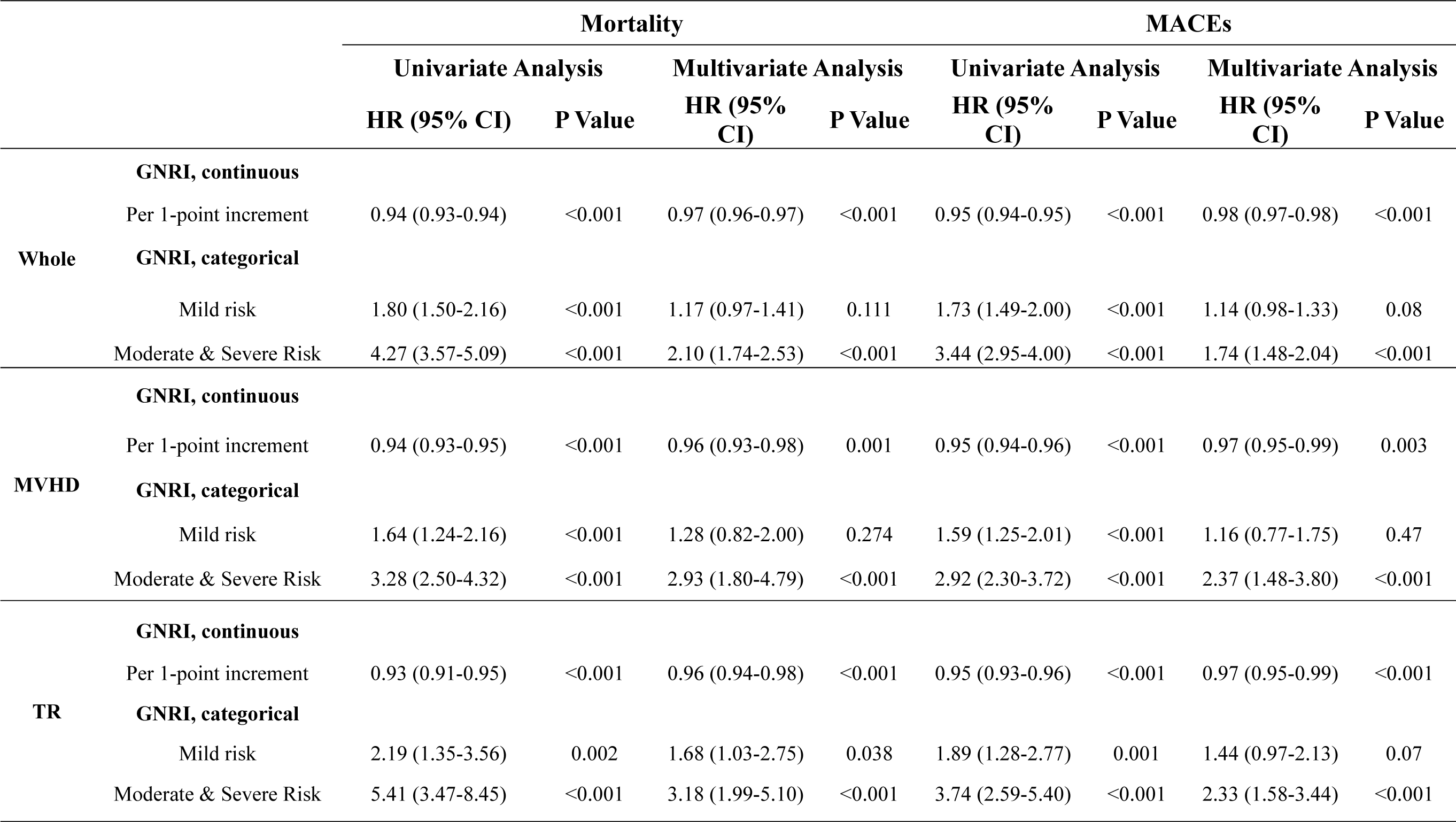
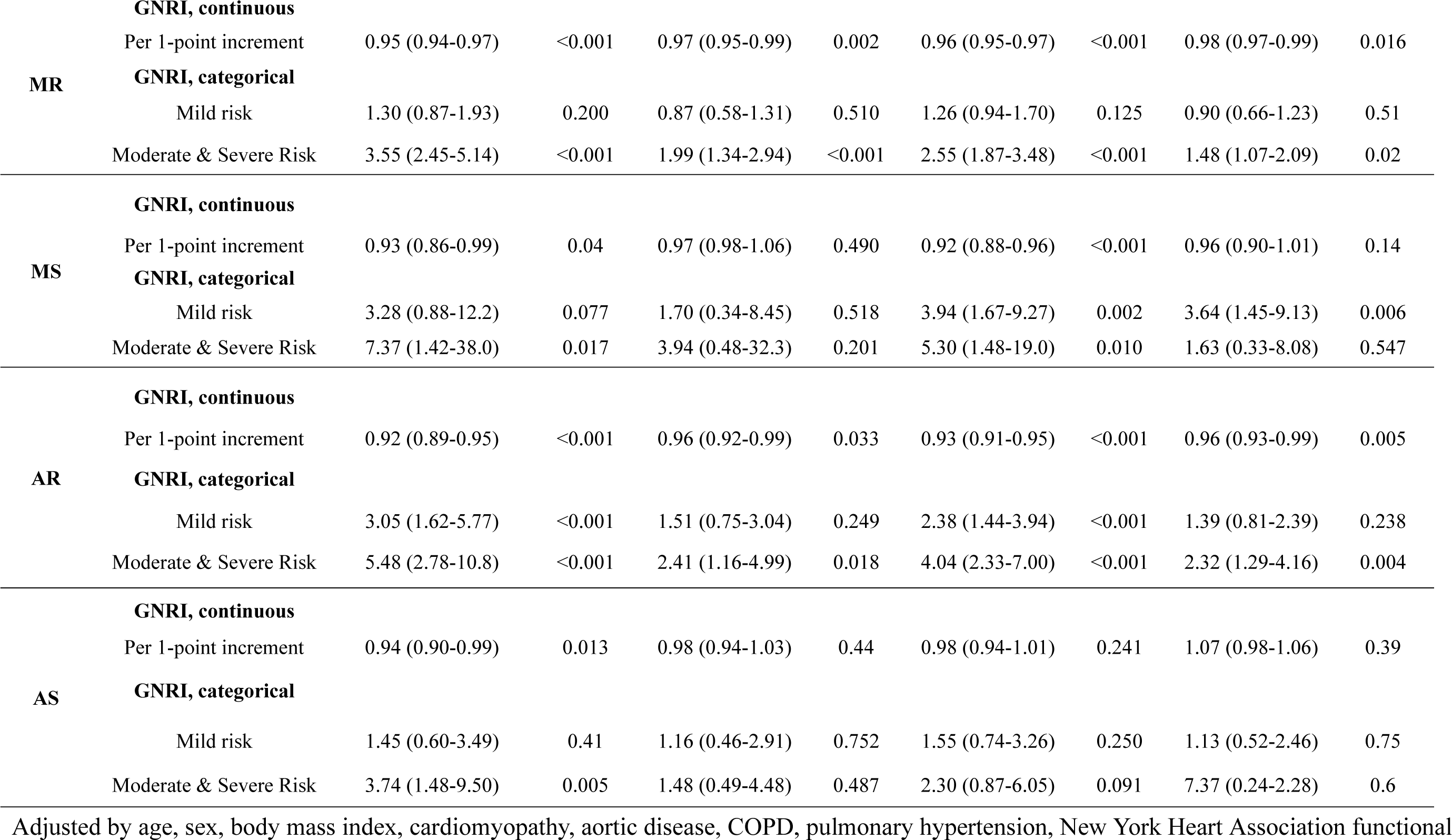
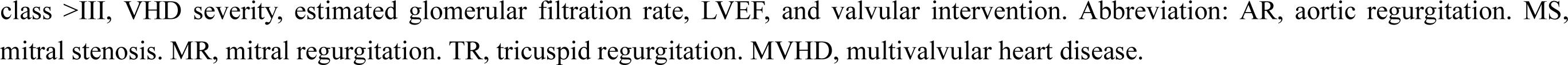
Cox Proportional Hazards Analyses of Malnutrition Index to Predict All-Cause Mortality and MACEs

### Stratification Analysis

Subgroup analyses were conducted to evaluate the interaction effects of several factors, including age, gender, NYHA class, etiology and severity of VHD, and intervention strategy, on the association between malnutrition severity and mortality, and no interaction with these covariates were found (Table S9). Consistency in the results was observed across diverse patient subgroups, such as young and middle-aged patients, individuals with normal cardiac function, moderate VHD, primary VHD, and those who underwent conservative medical treatment.

Patients were further divided into four groups based on their BMI (high, BMI≥24kg/m^2^; low, BMI<24kg/m^2^) and GNRI scores (high risk, <92; low risk, ≥92). Regardless of obesity status, VHD patients with high malnutritional risk had a more unfavorable prognosis compared to those with low malnutritional risk (GNRI<92 & BMI <24, HR, 3.61, 95% CI, 2.95-4.42, p<0.001; GNRI<92 & BMI≥24, HR, 4.16, 95% CI, 3.11-5.55, p<0.001)(Figure 5). The results were consistent across the VHD spectrum in subgroup analysis, except for AS and MS, where the sample sizes were relatively small (Figure S2). We performed further analyses that took into account the combined impact of NT-proBNP, LVEF, or EuroSCORE II and GNRI. The relationship with the outcome was stronger in individuals who had evidence of both biomarkers showing worse values (NT-proBNP≥1000 pg/ml, LVEF<50%, or EuroSCORE II ≥4%) and high malnutrition risk (GNRI<92) compared to those who had either biomarker showing worse values or high malnutrition risk alone (Figure 5).

**Figure 5.**
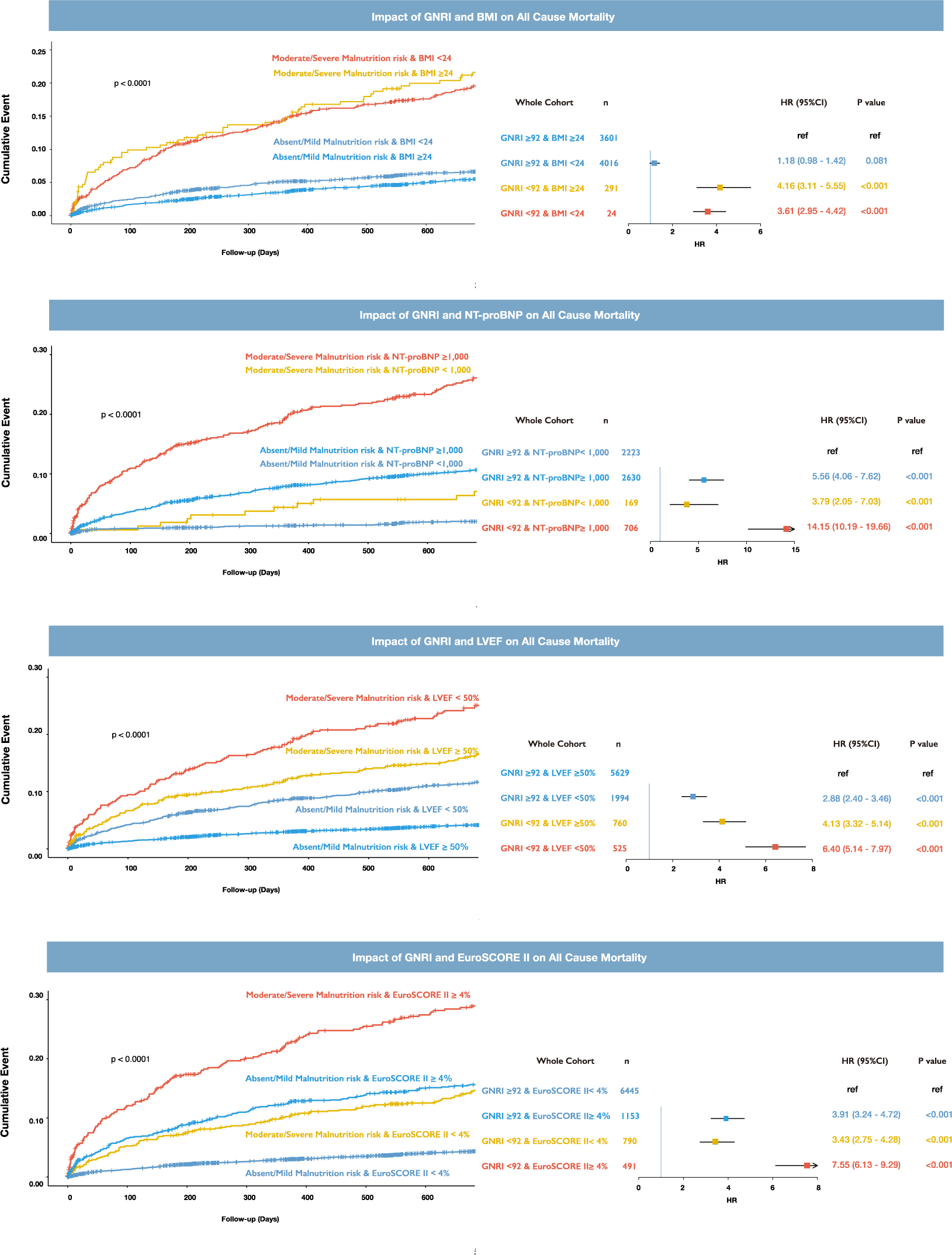
Malnutrition Degrees and Risk of Mortality. Abbreviation: AR, aortic regurgitation. MS, mitral stenosis. MR, mitral regurgitation. TR, tricuspid regurgitation. MVHD, multivalvular heart disease.

### Incremental Prognostic Value of GNRI score

Incorporating the GNRI score as a dichotomous variable (< vs. ≥92) enhanced risk prediction compared to EuroSCORE II alone (c-statistics: 0.73 vs. 0.71, p<0.001) (Table 3). The c-statistics for both primary and secondary outcomes were significantly greater when using the EuroSCORE II plus GNRI score (all p < 0.001) compared to using the EuroSCORE II alone (Table 3). Notably, the benefit of adding GNRI to the prediction algorithm was more substantial for MVHD, TR, AR, and MR, while it was only modest for AS and MS.

**Table 3.** Comparison of Risk Prediction Models With and Without GNRI for Overall Mortality

| Type of VHD | Model | C-statistic | LR test, *p* value | BIC |
| --- | --- | --- | --- | --- |
| Mortality |  |  |  |  |
| Overall | EuroSCORE II | 0.71 | <0.001 | 12,224 |
|  | EuroSCORE II+GNRI | 0.73 |  | 12,077 |
| MVHD | EuroSCORE II | 0.69 | <0.001 | 4645 |
|  | EuroSCORE II+GNRI | 0.70 |  | 4613 |
| TR | EuroSCORE II | 0.71 | <0.001 | 1644 |
|  | EuroSCORE II+GNRI | 0.74 |  | 1604 |
| MR | EuroSCORE II | 0.71 | <0.001 | 2277 |
|  | EuroSCORE II+GNRI | 0.73 |  | 2258 |
| MS | EuroSCORE II | 0.73 | 0.163 | 120 |
|  | EuroSCORE II+GNRI | 0.77 |  | 121 |
| AR | EuroSCORE II | 0.75 | <0.001 | 722 |
|  | EuroSCORE II+GNRI | 0.76 |  | 714 |
| AS | EuroSCORE II | 0.65 | 0.035 | 362 |
|  | EuroSCORE II+GNRI | 0.68 |  | 361 |
| MACE |  |  |  |  |
| Overall | EuroSCORE II | 0.69 | <0.001 | 17260 |
|  | EuroSCORE II+GNRI | 0.70 |  | 17139 |
| MVHD | EuroSCORE II | 0.66 | <0.001 | 6098 |
|  | EuroSCORE II+GNRI | 0.67 |  | 6068 |
| TR | EuroSCORE II | 0.72 | <0.001 | 2281 |
|  | EuroSCORE II+GNRI | 0.72 |  | 2251 |
| MR | EuroSCORE II | 0.70 | <0.001 | 3619 |
|  | EuroSCORE II+GNRI | 0.70 |  | 3609 |
| MS | EuroSCORE II | 0.68 | 0.285 | 271 |
|  | EuroSCORE II+GNRI | 0.66 |  | 273 |
| AR | EuroSCORE II | 0.69 | <0.001 | 1120 |
|  | EuroSCORE II+GNRI | 0.69 |  | 1112 |
| AS | EuroSCORE II | 0.63 | 0.256 | 454 |
|  | EuroSCORE II+GNRI | 0.65 |  | 457 |

Abbreviation: AR, aortic regurgitation. MS, mitral stenosis. MR, mitral regurgitation. TR, tricuspid regurgitation. MVHD, multivalvular heart disease. LR, likelihood ratio. BIC, bayesian information criterion.

## Discussion

Malnutrition risk, as determined by GNRI score, was prevalent in patients with various types of VHD. Moderate and severe malnutrition risk was an independent predictor of poor clinical outcomes, which was further validated across various VHD phenotypes, irrespective of BMI and cardiac function. Furthermore, the addition of GNRI score to the conventional risk scores (EuroSCORE II) improved the prediction of 2-year mortality. Our findings suggest that malnutrition is a common comorbidity in VHD, and GNRI score can help identify VHD patients who are at higher risk for adverse clinical events.

Malnutrition is a highly prevalent condition, particularly in patients with advanced disease stages.^17^ Several studies have investigated the prevalence of malnutrition in AS patients. Using the GNRI score, Seoudy et al. reported malnutrition rates of 35.2% and 38.4% in AS development and validation cohorts, respectively. ^4^ Goldfarb et al. found that 32.8% of severe AS patients were at risk of malnutrition, as assessed by the Mini Nutritional Assessment-Short Form.^3^ A recent study reported that all three nutritional screening tools (PNI, CONUT, and GNRI) showed good accuracy and predictive power in severe AS patients.^18^ In line with prior research, our study identified that 28.6% of AS patients were at risk of malnutrition. The differences in reported malnutrition prevalence among studies might be due to differences in AS severity, with previous studies involving patients with more advanced disease stages. Additionally, our study contributed to the understanding of malnutrition risk in other types of VHD. Notably, the prevalence of malnutrition risk was higher in patients with MVHD (50.0%), TR (48.5%), and MR (40.0%) compared to AS (28.6%). Previous studies have reported high rates of malnutrition risk in patients with TR or MR, with 94% of 86 TR patients and 74.4% of 892 MR patients found to be malnourished or at risk of malnutrition using different nutritional assessment tools.^19, 20^ There may be a higher risk of malnutrition among patients with regurgitation lesions in the mitral and tricuspid valve, as these lesions are potentially linked to increased rates of right-sided heart failure. Such patients often present with liver and gastrointestinal congestion, which can result in pressure overload on the hepatic sinusoids, leading to hepatocyte atrophy and apoptosis and impairing the liver’s ability to regulate metabolism.^21, 22^Moreover, venous congestion may elevate lymphatic pressure, increase mucosal permeability, and promote endotoxin absorption, creating a pro-inflammatory environment that may contribute to cardiac cachexia, a hallmark of advanced-stage VHD. ^23, 24^

Regarding the etiology of valve heart disease (VHD), malnutrition risk is relatively high in patients with degenerative (44.5%), functional (48.2%), and ischemic heart disease (47.7%) compared to patients with rheumatic heart disease (32.5%) and congenital heart disease (32.3%). The varying malnutrition risk in different etiologies of VHD may be related to the different mechanisms of malnutrition development in each type of VHD. The classification of malnutrition comprises three etiological subtypes: disease-related malnutrition driven by inflammation, disease-related malnutrition without inflammation, and malnutrition without disease.^17^ In degenerative valve disease, upregulation of chronic inflammatory cytokines causes sustained low-grade inflammation that affects the central nervous system and inhibits gastric emptying, thereby leading to inflammation-mediated malnutrition.^25^ Malnutrition in functional and ischemic valve disease is commonly caused by non-inflammatory systemic diseases, such as stroke and dementia, that can result in a decline in sensory functions (e.g., taste, smell, and vision) and difficulty swallowing. Malnutrition in rheumatic valve disease may be linked to inadequate nutrient intake caused by economic insufficiency, as the disease itself is associated with poverty and poor living conditions. ^26^As China’s economy has developed, the proportion of malnutrition caused by insufficient food sources has decreased, reducing the risk of malnutrition in rheumatic valve disease. The lower risk of malnutrition in congenital valve disease is mainly attributed to age-related factors, as the disease typically presents at a younger age and is less commonly accompanied by other systemic diseases.

Our study highlighted the importance of assessing malnutrition risk in VHD patients, as we found that worsening malnutrition status was linked to increased all-cause mortality, particularly in AR, MR, TR, and MVHD patients. Previous studies have suggested that a low GNRI score was associated with higher mortality risk in patients with severe AS, and that procedural nutritional status measured by the Mini Nutritional Assessment-Short Form was able to predict all-cause mortality in patients undergoing AVR.^3, 4^ However, in present study, the association between GNRI score and the prognosis of patients with AS was found to be close to null. This may be due to the relatively small number of AS patients included in our study, the less severe form of AS disease, and the shorter follow-up period, which limited the number of clinical outcomes and reduced the statistical power to detect clinically significant differences. Thus, to confirm our findings in AS patients, a longer follow-up period is necessary for this subgroup of patients.

In addition to being prevalent in underweight individuals, malnutrition risk was also found in overweight and obese patients with VHD. The malnutrition score was a stronger predictor of mortality than BMI, indicating that malnutrition was not solely linked to being underweight. Therefore, it is crucial to conduct nutritional assessments in obese patients with VHD. Although the GNRI score alone does not provide the exact prevalence of malnutrition, it is an easily calculated and objective index that enables clinicians to screen for malnutrition risk in VHD patients using routine biomarker measurements. Besides, the assessment of nutritional status, combined with the EuroSCORE II score, can improve risk stratification for VHD patients and aid in secondary prevention interventions. This approach can identify patients at risk of adverse events and help determine who may benefit from nutritional support. To facilitate the nutritional care process, a multidisciplinary approach involving physicians and dieticians should be adopted in cardiac rehabilitation, with early commencement of nutritional care during the initial VHD hospitalization.^27^

Our study had several limitations. Firstly, we were unable to compare our findings with other nutritional assessment tools, such as the Mini-Nutritional Assessment short-form, PNI, or CONUT scores for risk stratification, despite there is currently no consensus on assessment criteria or standard methods for assessing nutritional status in VHD patients. Therefore, confirmation of our results with other tools for nutritional risk assessment would be necessary. Secondly, the number of events in moderate to severe groups of AS and MS was relatively small, which may have limited our statistical power to detect clinically significant differences in adverse event rates in specific subsets. Thirdly, we could not assess how temporal changes in GNRI score may affect the association with adverse clinical outcomes. Fourthly, we could not capture the use of nutritional supplements prescribed by clinical providers, and therefore could not adjust them in regression models. Finally, as our study only included Chinese patients, the generalizability of our findings to other ethnicities needs confirmation.

## Conclusion

Our study highlighted the high prevalence of malnutrition risk among VHD patients, and demonstrated that those with poor nutritional status were at increased risk of all-cause mortality and MACE events. Malnutrition assessment is therefore crucial for identifying patients who are at elevated risk of adverse clinical outcomes and who may benefit from nutritional support. These findings provide a basis for future prospective randomized studies to evaluate the efficacy of nutritional interventions on outcomes in VHD patients.

## Data Availability

The data are available from the corresponding author upon reasonable request.

## Acknowledgments

This work was supported by the Chinese Academy of Medical Sciences Innovation Fund for Medical Sciences [2017-12M-3-002].

## Conflict of interest

none declared.

## Ethical guidelines statement

All participants provided written informed consent. Ethics approval was granted by the Human Ethics Committee of the Fuwai hospital. The study was performed in accordance with the ethical standards laid down in the 1964 Declaration of Helsinki and its later amendments.

## Notes

### Competing Interest Statement

The authors have declared no competing interest.

### Author Declarations

Human Ethics Committee of the Fuwai hospital

## Reference

1. Coffey S, Roberts-Thomson R, Brown A, Carapetis J, Chen M, Enriquez-Sarano M, Zühlke L and Prendergast BD. Global epidemiology of valvular heart disease. Nat Rev Cardiol. 2021;18:853–864.

2. Messika-Zeitoun D, Baumgartner H, Burwash IG, Vahanian A, Bax J, Pibarot P, Chan V, Leon M, Enriquez-Sarano M, Mesana T and Iung B. Unmet needs in valvular heart disease. Eur Heart J. 2023.

3. Goldfarb M, Lauck S, Webb JG, Asgar AW, Perrault LP, Piazza N, Martucci G, Lachapelle K, Noiseux N, Kim DH, Popma JJ, Lefèvre T, Labinaz M, Lamy A, Peterson MD, Arora RC, Morais JA, Morin JF, Rudski LG and Afilalo J. Malnutrition and Mortality in Frail and Non-Frail Older Adults Undergoing Aortic Valve Replacement. Circulation. 2018;138:2202–2211.

4. Seoudy H, Al-Kassou B, Shamekhi J, Sugiura A, Frank J, Saad M, Bramlage P, Seoudy AK, Puehler T, Lutter G, Schulte DM, Laudes M, Nickenig G, Frey N, Sinning JM and Frank D. Frailty in patients undergoing transcatheter aortic valve replacement: prognostic value of the Geriatric Nutritional Risk Index. J Cachexia Sarcopenia Muscle. 2021;12:577–585.

5. Zhang G, Pan Y, Zhang R, Wang M, Meng X, Li Z, Li H, Wang Y, Zhao X, Liu G and Wang Y. Prevalence and Prognostic Significance of Malnutrition Risk in Patients With Acute Ischemic Stroke: Results From the Third China National Stroke Registry. Stroke. 2022;53:111–119.

6. Minamisawa M, Seidelmann SB, Claggett B, Hegde SM, Shah AM, Desai AS, Lewis EF, Shah SJ, Sweitzer NK, Fang JC, Anand IS, O’Meara E, Rouleau JL, Pitt B and Solomon SD. Impact of Malnutrition Using Geriatric Nutritional Risk Index in Heart Failure With Preserved Ejection Fraction. JACC Heart Fail. 2019;7:664–675.

7. Raposeiras Roubín S, Abu Assi E, Cespón Fernandez M, Barreiro Pardal C, Lizancos Castro A, Parada JA, Pérez DD, Blanco Prieto S, Rossello X, Ibanez B and Íñiguez Romo A. Prevalence and Prognostic Significance of Malnutrition in Patients With Acute Coronary Syndrome. J Am Coll Cardiol. 2020;76:828–840.

8. Anzaki K, Kanda D, Ikeda Y, Takumi T, Tokushige A, Ohmure K, Sonoda T, Arikawa R and Ohishi M. Impact of Malnutrition on Prognosis and Coronary Artery Calcification in Patients with Stable Coronary Artery Disease. Curr Probl Cardiol. 2022:101185.

9. González Ferreiro R, López Otero D, Álvarez Rodríguez L, Otero García Ó, Pérez Poza M, Antúnez Muiños PJ, Cacho Antonio C, López Pais J, Juskowa M, Cid Álvarez AB, Trillo Nouche R, Sanmartín Pena XC, Sánchez Fernández PL, Cruz-González I and González Juanatey JR. Prognostic Impact of Change in Nutritional Risk on Mortality and Heart Failure After Transcatheter Aortic Valve Replacement. Circ Cardiovasc Interv. 2021;14:e009342.

10. Vahanian A, Beyersdorf F, Praz F, Milojevic M, Baldus S, Bauersachs J, Capodanno D, Conradi L, De Bonis M, De Paulis R, Delgado V, Freemantle N, Gilard M, Haugaa KH, Jeppsson A, Jüni P, Pierard L, Prendergast BD, Sádaba JR, Tribouilloy C and Wojakowski W. 2021 ESC/EACTS Guidelines for the management of valvular heart disease. Eur Heart J. 2022;43:561–632.

11. Nashef SA, Roques F, Sharples LD, Nilsson J, Smith C, Goldstone AR and Lockowandt U. EuroSCORE II. Eur J Cardiothorac Surg. 2012;41:734–44; discussion 744-5.

12. von Elm E, Altman DG, Egger M, Pocock SJ, Gøtzsche PC and Vandenbroucke JP. The Strengthening the Reporting of Observational Studies in Epidemiology (STROBE) statement: guidelines for reporting observational studies. Lancet. 2007;370:1453–7.

13. Lang RM, Badano LP, Mor-Avi V, Afilalo J, Armstrong A, Ernande L, Flachskampf FA, Foster E, Goldstein SA, Kuznetsova T, Lancellotti P, Muraru D, Picard MH, Rietzschel ER, Rudski L, Spencer KT, Tsang W and Voigt JU. Recommendations for cardiac chamber quantification by echocardiography in adults: an update from the American Society of Echocardiography and the European Association of Cardiovascular Imaging. J Am Soc Echocardiogr. 2015;28:1–39.e14.

14. Lv J, Ye Y, Li Z, Zhang B, Liu Q, Zhao Q, Zhao Z, Wang W, Zhang H, Duan Z, Wang B, Yu Z, Guo S, Zhao Y, Gao R, Xu H and Wu Y. Prognostic Value of Modified Model for End-Stage Liver Disease Scores in Patients With Significant Tricuspid Regurgitation. Eur Heart J Qual Care Clin Outcomes. 2022.

15. Bouillanne O, Morineau G, Dupont C, Coulombel I, Vincent JP, Nicolis I, Benazeth S, Cynober L and Aussel C. Geriatric Nutritional Risk Index: a new index for evaluating at-risk elderly medical patients. Am J Clin Nutr. 2005;82:777–83.

16. Pan XF, Wang L and Pan A. Epidemiology and determinants of obesity in China. Lancet Diabetes Endocrinol. 2021;9:373–392.

17. Dent E, Wright ORL, Woo J and Hoogendijk EO. Malnutrition in older adults. Lancet. 2023;401:951–966.

18. Ishizu K, Shirai S, Tashiro H, Kitano K, Tabata H, Nakamura M, Morofuji T, Murakami N, Morinaga T, Hayashi M, Isotani A, Arai Y, Ohno N, Kakumoto S and Ando K. Prevalence and Prognostic Significance of Malnutrition in Older Japanese Adults at High Surgical Risk Undergoing Transcatheter Aortic Valve Implantation. J Am Heart Assoc. 2022;11:e026294.

19. Caneiro-Queija B, Raposeiras-Roubin S, Adamo M, Freixa X, Arzamendi D, Benito-González T, Montefusco A, Pascual I, Nombela-Franco L, Rodes-Cabau J, Shuvy M, Portolés-Hernández A, Godino C, Haberman D, Lupi L, Regueiro A, Li CH, Fernández-Vázquez F, Frea S, Avanzas P, Tirado-Conte G, Paradis JM, Peretz A, Moñivas V, Baz JA, Galasso M, Branca L, Sanchís L, Asmarats L, Garrote-Coloma C, Angelini F, León V, de Agustín JA, Alperi A, Beeri R, Maccagni G, Sabaté M, Fernández-Peregrina E, Gualis J, Bocchino PP, Curello S, Íñiguez-Romo A and Estévez-Loureiro R. Prognostic Impact of Nutritional Status After Transcatheter Edge-to-Edge Mitral Valve Repair: The MIVNUT Registry. J Am Heart Assoc. 2022;11:e023121.

20. Besler C, Unterhuber M, Rommel KP, Unger E, Hartung P, von Roeder M, Noack T, Zachäus M, Halm U, Borger M, Desch S, Thiele H and Lurz P. Nutritional status in tricuspid regurgitation: implications of transcatheter repair. Eur J Heart Fail. 2020;22:1826–1836.

21. Gore RM, Mathieu DG, White EM, Ghahremani GG, Panella JS and Rochester D. Passive hepatic congestion: cross-sectional imaging features. AJR Am J Roentgenol. 1994;162:71–5.

22. Samsky MD, Patel CB, DeWald TA, Smith AD, Felker GM, Rogers JG and Hernandez AF. Cardiohepatic interactions in heart failure: an overview and clinical implications. J Am Coll Cardiol. 2013;61:2397–2405.

23. Colombo PC, Ganda A, Lin J, Onat D, Harxhi A, Iyasere JE, Uriel N and Cotter G. Inflammatory activation: cardiac, renal, and cardio-renal interactions in patients with the cardiorenal syndrome. Heart Fail Rev. 2012;17:177–90.

24. Valentova M, von Haehling S, Bauditz J, Doehner W, Ebner N, Bekfani T, Elsner S, Sliziuk V, Scherbakov N, Murín J, Anker SD and Sandek A. Intestinal congestion and right ventricular dysfunction: a link with appetite loss, inflammation, and cachexia in chronic heart failure. Eur Heart J. 2016;37:1684–91.

25. Galante A, Pietroiusti A, Vellini M, Piccolo P, Possati G, De Bonis M, Grillo RL, Fontana C and Favalli C. C-reactive protein is increased in patients with degenerative aortic valvular stenosis. J Am Coll Cardiol. 2001;38:1078–82.

26. Marijon E, Mirabel M, Celermajer DS and Jouven X. Rheumatic heart disease. Lancet. 2012;379:953–964.

27. Sun YP and O’Gara PT. Beyond Frailty and Conventional Risk Scores. Circulation. 2018;138:2212–2215.

